# Short-term Care Burden After Left Ventricular Assist Device Implantation and Heart Transplant in the United States

**DOI:** 10.1101/2025.05.02.25326911

**Authors:** Balaphanidhar Mogga, Ashwin Pillai, Karanrajsinh Raol, William L. Baker, David A. Baran, Prathyusha Muddluru, Zeina Jedeon, Sreenivas Sagili, Abhishek Jaiswal

**Author notes:** <u>Correspondence:</u> Abhishek Jaiswal, MD, Hartford HealthCare Heart and Vascular Institute, Hartford Hospital, 85 Jefferson Street, Suite-JB208, Hartford, CT 06102, USA. The authors report no conflicts to disclose. Balaphanidhar Mogga, Karanrajsinh Raol, and Abhishek Jaiswal designed the study; all authors participated in writing this paper.

## Abstract

**Background:** Heart transplant (HT) and left ventricular assist devices (LVADs) are treatment options for advanced heart failure refractory to standard therapy. Historically, LVADs have been used as either destination therapy or a bridge to transplant. However, recent changes to the organ allocation system have deprioritized patients on LVADs as transplant recipients, leading to divisive views on the role of an LVAD. Comparative short term care burden with each modality remains unclear.

**Objectives:** To describe and compare characteristics, outcomes and cost burden during index hospitalizations, 30-day and 90-day readmissions associated with heart transplants (HT) and durable left ventricular assist devices (LVAD) from a large, national administrative database.

**Methods:** A review of the Nationwide Readmissions Database (NRD) from 2018-2021 describing a cohort identified using ICD-10-CM procedure codes: 02YA0Z0 and 02HA0QZ.

**Results:** We identified 27,308 index hospital admissions; 52.4% received LVADs and the remainder received HT. Compared to HT recipients LVAD recipients were older and more likely male, of lower socio-economic status, had longer index hospital stays, and more expensive index hospitalizations. A higher proportion of LVAD recipients required rehospitalizations in 30 days. The most frequent causes of re-hospitalization in the LVAD group at both 30 and 90 days were heart failure, device complications, and gastrointestinal bleeding. The most frequent causes in the HT group at 30 and 90 days were transplant complications, renal dysfunction, and sepsis (at 90 days only). 30 and 90-day rehospitalization costs were greater in the HT group.

**Conclusions:** Both LVAD and HT had comparable burdens on resource and short-term rehospitalization risk.

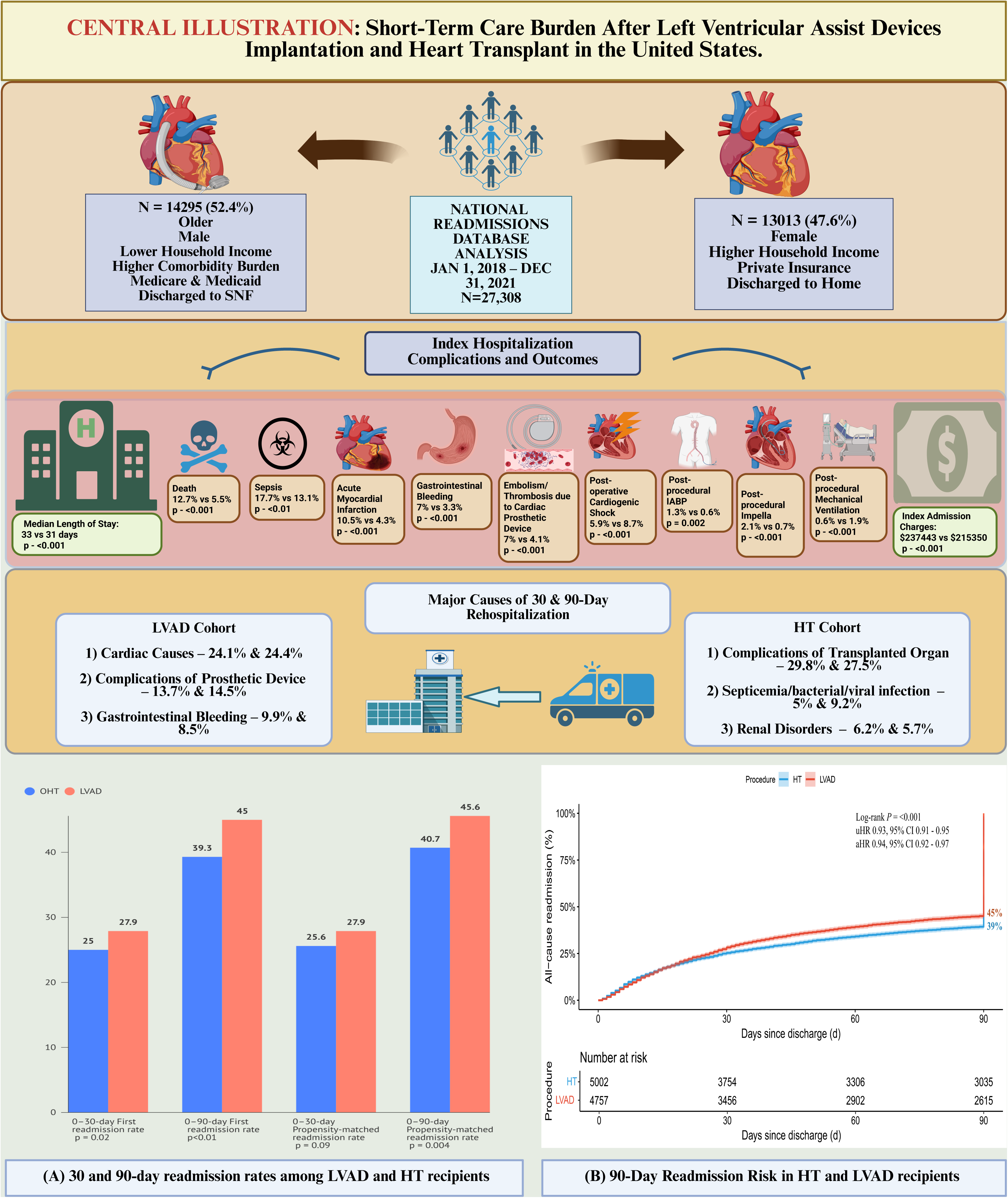

## Introduction

Heart Transplantation (HT) and mechanical circulatory support with a durable left ventricular assist device (LVAD) both improve survival in eligible patients with heart failure who do not respond to medical therapy. Durability and perceived fewer complications enhance the value of heart transplants compared to durable LVADs. Usually, an LVAD is offered until HT can be performed or as a destination therapy when HT no longer remains a viable therapeutic option. However, the HeartMate 3 (HM3), the latest fully magnetically levitated, frictionless LVAD pump, is engineered with wide blood flow pathways and pulsatility in the pump due to programmatic fixed rotor speed changes, has reduced LVAD-related complications and improved long-term survival compared to older devices, rivaling HT.^1,2^

The information above is also increasingly relevant, as the latest organ allocation system for HT in adults in the United States has reduced the priority for patients with an LVAD awaiting HT. ^3^ As a result, the use of durable LVADs has decreased vis-à-vis the increase in the use of temporary devices as a bridge to HT. These evolving perspectives have led to differing opinions on the appropriate timing and circumstances for LVAD or HT. With comparable 5-year survivals, the burden of cost, resource utilization, and readmissions are of growing interest to patients and providers alike while weighing both options.

While the Centers for Medicare and Medicaid Services monitor 30-day all-cause readmission rates for heart failure, acute myocardial infarction, and other conditions to maintain quality measures, such a process is not in place for patients receiving LVAD or HT. Hence, there is a lack of representative data on rehospitalizations, length of stay (LOS), healthcare costs, and resource utilization for HT patients and those with the HM3 LVAD therapy in real-world clinical care settings.

Therefore, we analyzed a nationwide database for patterns of use, outcomes, and resource utilization after HT and LVAD implantation, focusing on the years following approval of the latest generation HM3 LVAD.

## Methods

### Data Source

The Nationwide Readmissions Database (NRD) was developed by the Healthcare Cost and Utilization Project (HCUP) of the United States Agency for Healthcare Research and Quality (AHRQ).^4^ Sponsored by the AHRQ within the HCUP, the NRD draws from state inpatient databases and employs reliable patient linkage numbers to facilitate tracking of individual readmissions across various hospitals. It is the largest, all-payer-inclusive inpatient database that is publicly available. Each patient is assigned a unique identifier code (NRD_VistLink) for tracing readmissions within a calendar year. The NRD days-to-event variable captures readmissions within a calendar year but not across different years. Our study was exempt from IRB approval because it utilized publicly available, de-identified data. The data, analytic methods, and study materials are available through the HCUP website and may be used to reproduce the results.

### Study Cohort

The study population included new recipients of HT or LVAD using the International Classification of Diseases, 10th edition Clinical Modification Procedure Codes (ICD-10-CM procedure codes: 02YA0Z0 and 02HA0QZ) from January 1^st^, 2018, through December 31^st^, 2021. We selected this recent time frame to limit the influence of older LVAD technology. We excluded patients aged <18 years, who had missing information on the corresponding diagnosis-related group (DRG) codes at discharge, missing LOS, and those who had both HT and LVAD during the index hospitalization. We excluded patients admitted in December for the 30-day readmission cohort (30-DRC) to ensure a 30-day follow-up within the same calendar year. Similarly, for the 90-day readmission cohort (90-DRC), we excluded patients admitted in October, November, and December to ensure a 90-day follow-up within the same year.

We analyzed patient characteristics including age, gender, discharge disposition, income quartiles, hospital bed size and ownership status, teaching hospital status, primary payment methods, household income, and comorbid conditions. In addition, we also analyzed characteristics of index admission, such as the cost, LOS, average number of procedures, and pre-procedural hemodynamic /respiratory support.

### Outcome Variables

The primary outcomes were 30-day and 90-day readmission rates after discharge from the index hospitalization for LVAD or HT. After determining the target population, [NRD_VisitLink] and [NRD_DaysToEvent] were used to find subsequent readmissions and days for each readmission. We followed the methodology described by HCUP for readmission analysis.^5^

Secondary outcomes included resource utilization, LOS, and cost of hospitalization during the index hospitalization as well as readmissions, index hospitalization complications, index hospitalization dispositions, and reasons for readmissions. Reasons for readmissions were ascertained using primary diagnosis categories. Patients transferred to other hospitals were not considered in the readmission analysis.

To derive cost estimates for the index admission and readmissions, we recalculated the total hospital charges using a hospital-specific cost-to-charge ratio provided by the NRD. As the charges for each admission typically far exceed the actual care costs due to discrepancies driven by reimbursement policies, this method ensured our financial analysis reflects a more accurate representation of the actual costs associated with patient care. The cost information was adjusted for inflation and reported as 2020 US dollars using the Bureau of Labor Statistics CPI inflation calculator (Bureau of Labor Statistics. CPI Inflation Calculator. Accessed November 29, 2023. https://data.bls.gov/cgi-bin/cpicalc.pl). We also performed cost analysis as related to age group, sex, primary insurance payer, hospital control, hospital bed size, and median household income.

### Statistical Analyses

STATA svy command was implemented to handle survey data, ensuring that findings are representative of a national scale using weighted data, enhancing the generalizability of the conclusions. Descriptive analysis was performed to summarize patient characteristics. Categorical variables were reported as counts and percentages and compared using Pearson’s chi-square test. Continuous variables were summarized as medians with interquartile ranges (IQR) and compared using the Mann-Whitney U test. Hierarchical or multilevel models, which consider the impact of nesting (e.g., patient-level effects within hospital-level effects), were employed for multivariable analyses. In these models, a unique hospital identification number was included as a random effect, resulting in a two-level model.^6^ This incorporation of random effects allowed for the accommodation of variability at both the patient and hospital levels. We used mixed-effects logistic regression to evaluate the association between total cost and 30-day readmission, adjusting for patient, hospital, and clinical covariates, and determined Odds Ratios (OR) with 95% confidence intervals (CI). In all multivariable models, interaction terms such as age and payer were assessed and retained only if they were statistically significant (p < 0.05). A backward elimination approach was applied to refine the model, removing nonsignificant variables while ensuring the primary predictor’s stability. The final parsimonious model included both fixed effects and random effects at the hospital level, offering robust and interpretable estimates for our research objectives. A complete list of variables is listed in **Supplement Table 1.** Additionally, we analyzed time-to-first readmission using the Kaplan-Meier plot.

A secondary analysis was performed using a propensity score matching methodology to match patients with HT to those with LVAD in a 1:1 ratio. Each case was matched to a control using the nearest-neighbor technique with a caliper width of 0.02 without replacement. The balance of covariates before and after propensity score matching was assessed using the absolute standardized mean difference (**Supplement Figures 1-4**). The propensity score was calculated based on the same variables included in the multivariable logistic regression model (**Supplement Table 1**). Since missing values were limited to <1% for all included outcomes and variables, all analyses were performed using complete cases. For all analyses, a 2-tailed P-value of < 0.05 was considered statistically significant. Analyses were performed using Stata Statistical Software, Version 18 (StataCorp LLC, located in College Station, Texas, USA).

## Results

### Baseline characteristics of the Index study population

During the study period, 14,369 heart transplants and LVAD implantations accounted for 27,308 weighted index hospital admissions; LVAD was implanted in 52.4%. The change in donor allocation for HT decreased annual LVAD implantation (**Figure 1**).

**Figure 1:**
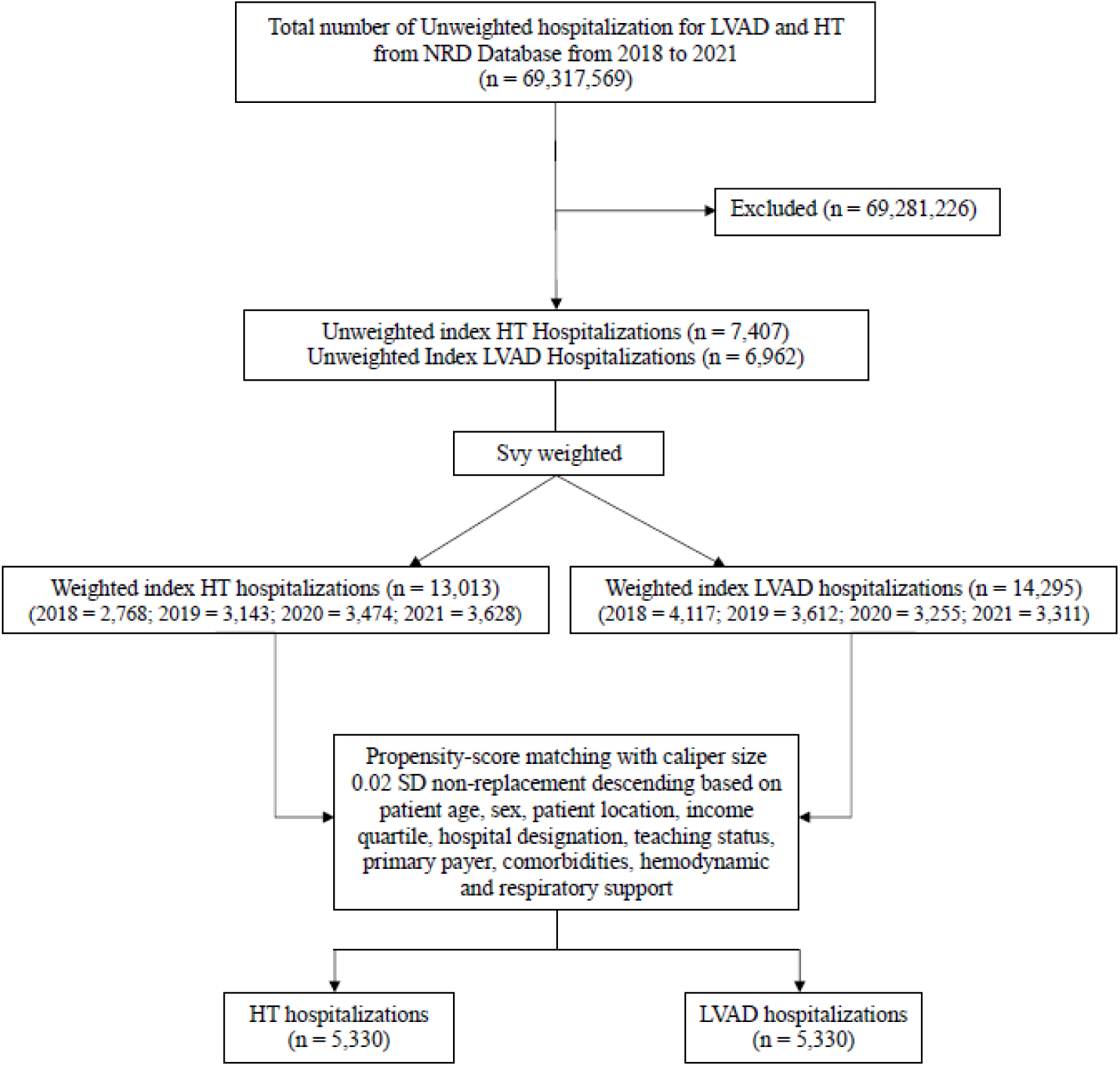
Study population.

**Table 1** displays a comparative analysis of baseline characteristics of the index hospitalization study cohort. Compared to HT, implantation of LVADs was more likely in older individuals (59 vs. 56 years, p<0.001), males (78% vs 72.6% p<0.001), individuals from the lowest-income quartile (30.9% vs 23%, p <0.001) and with Medicare as their primary payer (47.6 % vs 35.3 %, p <0.001) and was associated with a longer total LOS (33 vs 31 days, p<0.001) and post-procedural LOS (21 vs 17 days, p < 0.001).

**Table 1:**
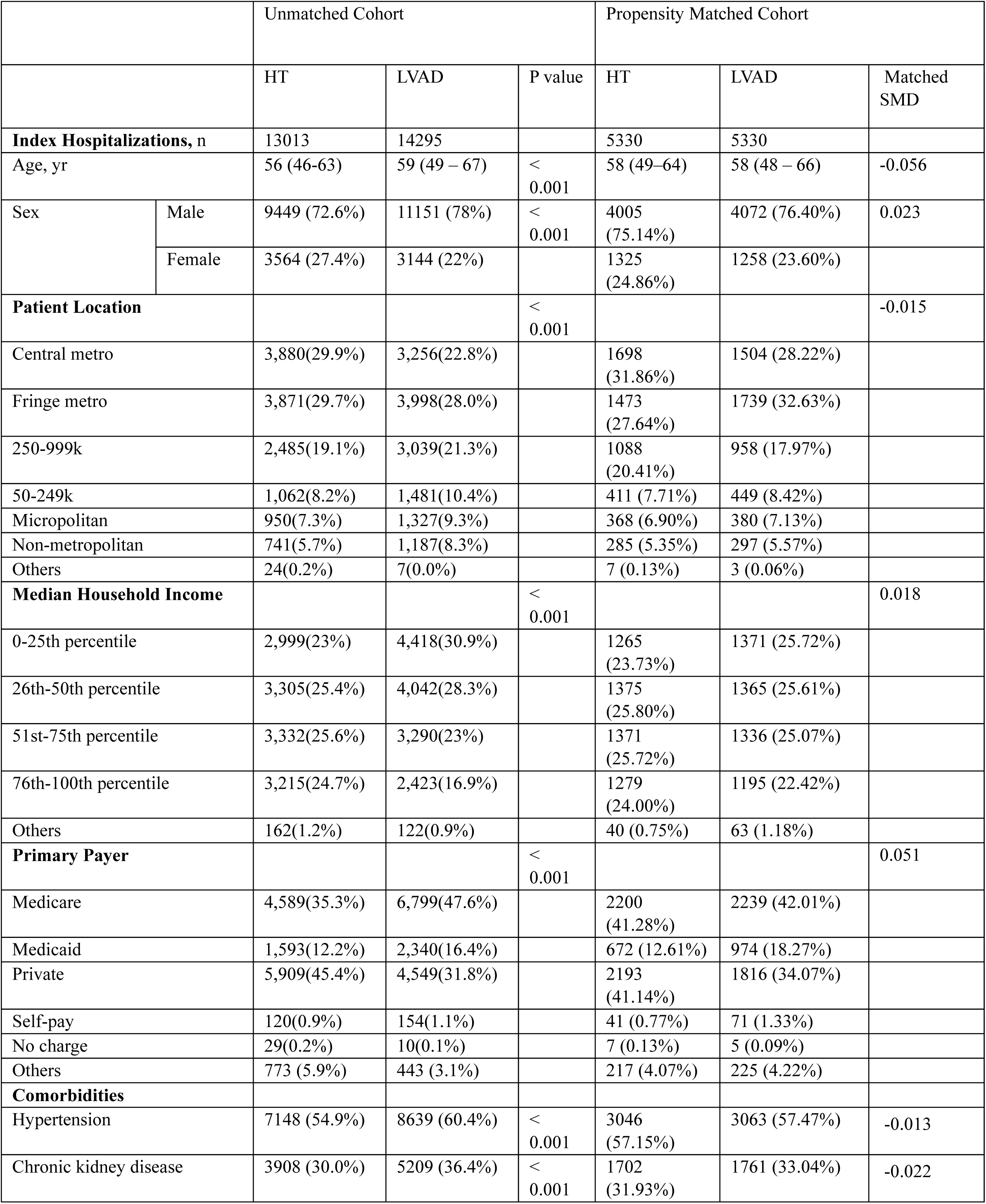
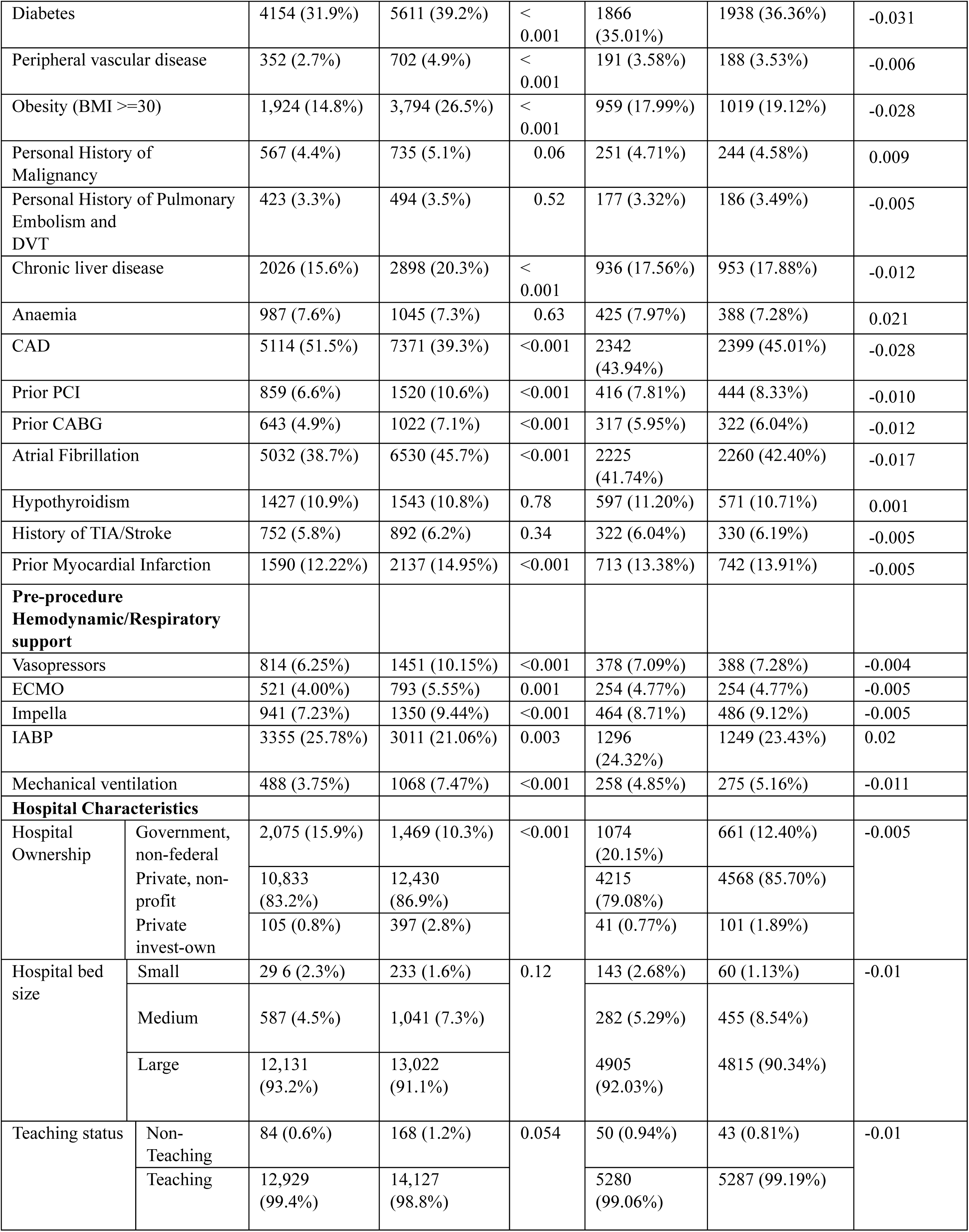
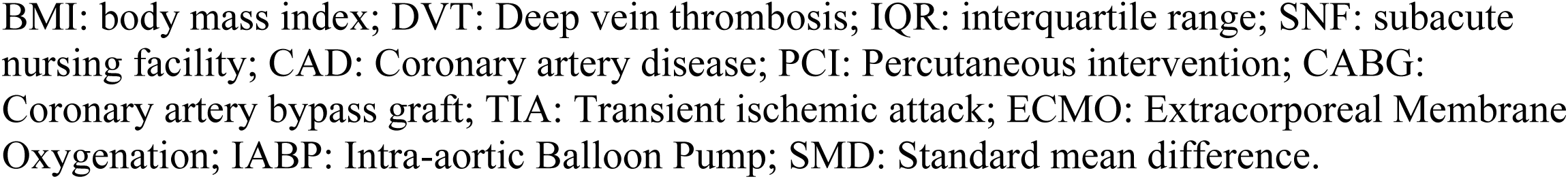
Baseline Characteristics of Index Hospitalization Study Group.

|  |  | Unmatched Cohort |  |  | Propensity Matched Cohort |  |  |
| --- | --- | --- | --- | --- | --- | --- | --- |
|  |  | HT | LVAD | P value | HT | LVAD | Matched SMD |
| **Index Hospitalizations, n** |  | 13013 | 14295 |  | 5330 | 5330 |  |
| Age, yr |  | 56 (46-63) | 59 (49 – 67) | < 0.001 | 58 (49–64) | 58 (48 – 66) | -0.056 |
| Sex | Male | 9449 (72.6%) | 11151 (78%) | < 0.001 | 4005 (75.14%) | 4072 (76.40%) | 0.023 |
|  | Female | 3564 (27.4%) | 3144 (22%) |  | 1325 (24.86%) | 1258 (23.60%) |  |
| **Patient Location** |  |  |  | < 0.001 |  |  | -0.015 |
| Central metro |  | 3,880(29.9%) | 3,256(22.8%) |  | 1698 (31.86%) | 1504 (28.22%) |  |
| Fringe metro |  | 3,871(29.7%) | 3,998(28.0%) |  | 1473 (27.64%) | 1739 (32.63%) |  |
| 250-999k |  | 2,485(19.1%) | 3,039(21.3%) |  | 1088 (20.41%) | 958 (17.97%) |  |
| 50-249k |  | 1,062(8.2%) | 1,481(10.4%) |  | 411 (7.71%) | 449 (8.42%) |  |
| Micropolitan |  | 950(7.3%) | 1,327(9.3%) |  | 368 (6.90%) | 380 (7.13%) |  |
| Non-metropolitan |  | 741(5.7%) | 1,187(8.3%) |  | 285 (5.35%) | 297 (5.57%) |  |
| Others |  | 24(0.2%) | 7(0.0%) |  | 7 (0.13%) | 3 (0.06%) |  |
| **Median Household Income** |  |  |  | < 0.001 |  |  | 0.018 |
| 0-25th percentile |  | 2,999(23%) | 4,418(30.9%) |  | 1265 (23.73%) | 1371 (25.72%) |  |
| 26th-50th percentile |  | 3,305(25.4%) | 4,042(28.3%) |  | 1375 (25.80%) | 1365 (25.61%) |  |
| 51st-75th percentile |  | 3,332(25.6%) | 3,290(23%) |  | 1371 (25.72%) | 1336 (25.07%) |  |
| 76th-100th percentile |  | 3,215(24.7%) | 2,423(16.9%) |  | 1279 (24.00%) | 1195 (22.42%) |  |
| Others |  | 162(1.2%) | 122(0.9%) |  | 40 (0.75%) | 63 (1.18%) |  |
| **Primary Payer** |  |  |  | < 0.001 |  |  | 0.051 |
| Medicare |  | 4,589(35.3%) | 6,799(47.6%) |  | 2200 (41.28%) | 2239 (42.01%) |  |
| Medicaid |  | 1,593(12.2%) | 2,340(16.4%) |  | 672 (12.61%) | 974 (18.27%) |  |
| Private |  | 5,909(45.4%) | 4,549(31.8%) |  | 2193 (41.14%) | 1816 (34.07%) |  |
| Self-pay |  | 120(0.9%) | 154(1.1%) |  | 41 (0.77%) | 71 (1.33%) |  |
| No charge |  | 29(0.2%) | 10(0.1%) |  | 7 (0.13%) | 5 (0.09%) |  |
| Others |  | 773 (5.9%) | 443 (3.1%) |  | 217 (4.07%) | 225 (4.22%) |  |
| **Comorbidities** |  |  |  |  |  |  |  |
| Hypertension |  | 7148 (54.9%) | 8639 (60.4%) | < 0.001 | 3046 (57.15%) | 3063 (57.47%) | -0.013 |
| Chronic kidney disease |  | 3908 (30.0%) | 5209 (36.4%) | < 0.001 | 1702 (31.93%) | 1761 (33.04%) | -0.022 |
| Diabetes |  | 4154 (31.9%) | 5611 (39.2%) | < 0.001 | 1866 (35.01%) | 1938 (36.36%) | -0.031 |
| Peripheral vascular disease |  | 352 (2.7%) | 702 (4.9%) | < 0.001 | 191 (3.58%) | 188 (3.53%) | -0.006 |
| Obesity (BMI >=30) |  | 1,924 (14.8%) | 3,794 (26.5%) | < 0.001 | 959 (17.99%) | 1019 (19.12%) | -0.028 |
| Personal History of Malignancy |  | 567 (4.4%) | 735 (5.1%) | 0.06 | 251 (4.71%) | 244 (4.58%) | 0.009 |
| Personal History of Pulmonary Embolism and DVT |  | 423 (3.3%) | 494 (3.5%) | 0.52 | 177 (3.32%) | 186 (3.49%) | -0.005 |
| Chronic liver disease |  | 2026 (15.6%) | 2898 (20.3%) | < 0.001 | 936 (17.56%) | 953 (17.88%) | -0.012 |
| Anaemia |  | 987 (7.6%) | 1045 (7.3%) | 0.63 | 425 (7.97%) | 388 (7.28%) | 0.021 |
| CAD |  | 5114 (51.5%) | 7371 (39.3%) | <0.001 | 2342 (43.94%) | 2399 (45.01%) | -0.028 |
| Prior PCI |  | 859 (6.6%) | 1520 (10.6%) | <0.001 | 416 (7.81%) | 444 (8.33%) | -0.010 |
| Prior CABG |  | 643 (4.9%) | 1022 (7.1%) | <0.001 | 317 (5.95%) | 322 (6.04%) | -0.012 |
| Atrial Fibrillation |  | 5032 (38.7%) | 6530 (45.7%) | <0.001 | 2225 (41.74%) | 2260 (42.40%) | -0.017 |
| Hypothyroidism |  | 1427 (10.9%) | 1543 (10.8%) | 0.78 | 597 (11.20%) | 571 (10.71%) | 0.001 |
| History of TIA/Stroke |  | 752 (5.8%) | 892 (6.2%) | 0.34 | 322 (6.04%) | 330 (6.19%) | -0.005 |
| Prior Myocardial Infarction |  | 1590 (12.22%) | 2137 (14.95%) | <0.001 | 713 (13.38%) | 742 (13.91%) | -0.005 |
| **Pre-procedure Hemodynamic/Respiratory support** |  |  |  |  |  |  |  |
| Vasopressors |  | 814 (6.25%) | 1451 (10.15%) | <0.001 | 378 (7.09%) | 388 (7.28%) | -0.004 |
| ECMO |  | 521 (4.00%) | 793 (5.55%) | 0.001 | 254 (4.77%) | 254 (4.77%) | -0.005 |
| Impella |  | 941 (7.23%) | 1350 (9.44%) | <0.001 | 464 (8.71%) | 486 (9.12%) | -0.005 |
| IABP |  | 3355 (25.78%) | 3011 (21.06%) | 0.003 | 1296 (24.32%) | 1249 (23.43%) | 0.02 |
| Mechanical ventilation |  | 488 (3.75%) | 1068 (7.47%) | <0.001 | 258 (4.85%) | 275 (5.16%) | -0.011 |
| **Hospital Characteristics** |  |  |  |  |  |  |  |
| Hospital Ownership | Government, non-federal | 2,075 (15.9%) | 1,469 (10.3%) | <0.001 | 1074 (20.15%) | 661 (12.40%) | -0.005 |
|  | Private, non-profit | 10,833 (83.2%) | 12,430 (86.9%) |  | 4215 (79.08%) | 4568 (85.70%) |  |
|  | Private invest-own | 105 (0.8%) | 397 (2.8%) |  | 41 (0.77%) | 101 (1.89%) |  |
| Hospital bed size | Small | 29 6 (2.3%) | 233 (1.6%) | 0.12 | 143 (2.68%) | 60 (1.13%) | -0.01 |
|  | Medium | 587 (4.5%) | 1,041 (7.3%) |  | 282 (5.29%) | 455 (8.54%) |  |
|  | Large | 12,131 (93.2%) | 13,022 (91.1%) |  | 4905 (92.03%) | 4815 (90.34%) |  |
| Teaching status | Non-Teaching | 84 (0.6%) | 168 (1.2%) | 0.054 | 50 (0.94%) | 43 (0.81%) | -0.01 |
|  | Teaching | 12,929 (99.4%) | 14,127 (98.8%) |  | 5280 (99.06%) | 5287 (99.19%) |  |

BMI: body mass index; DVT: Deep vein thrombosis; IQR: interquartile range; SNF: subacute nursing facility; CAD: Coronary artery disease; PCI: Percutaneous intervention; CABG: Coronary artery bypass graft; TIA: Transient ischemic attack; ECMO: Extracorporeal Membrane Oxygenation; IABP: Intra-aortic Balloon Pump; SMD: Standard mean difference.

### Complications and resource utilization

Major complications during index hospitalizations are summarized in **Figure 2 & Supplement Figure 5**. Compared with receiving an LVAD, receiving HT was associated with significantly lower in-hospital mortality (5.5% vs 12.7%; OR 0.45, 95% CI 0.38 to 0.54, p <0.001), acute gastrointestinal bleeding (3.3% vs 7%; OR 0.50, 95% CI 0.41 to 0.61, p <0.001), thromboembolic complications attributable to cardiac prosthetic devices (4.1% vs 7%; OR 0.67, 95% CI 0.54 to 0.83, p = 0.002), and acute myocardial infarction (4.3% vs 10.5%; OR 0.42, 95% CI 0.35 to 0.50, p <0.001).

**Figure 2:**
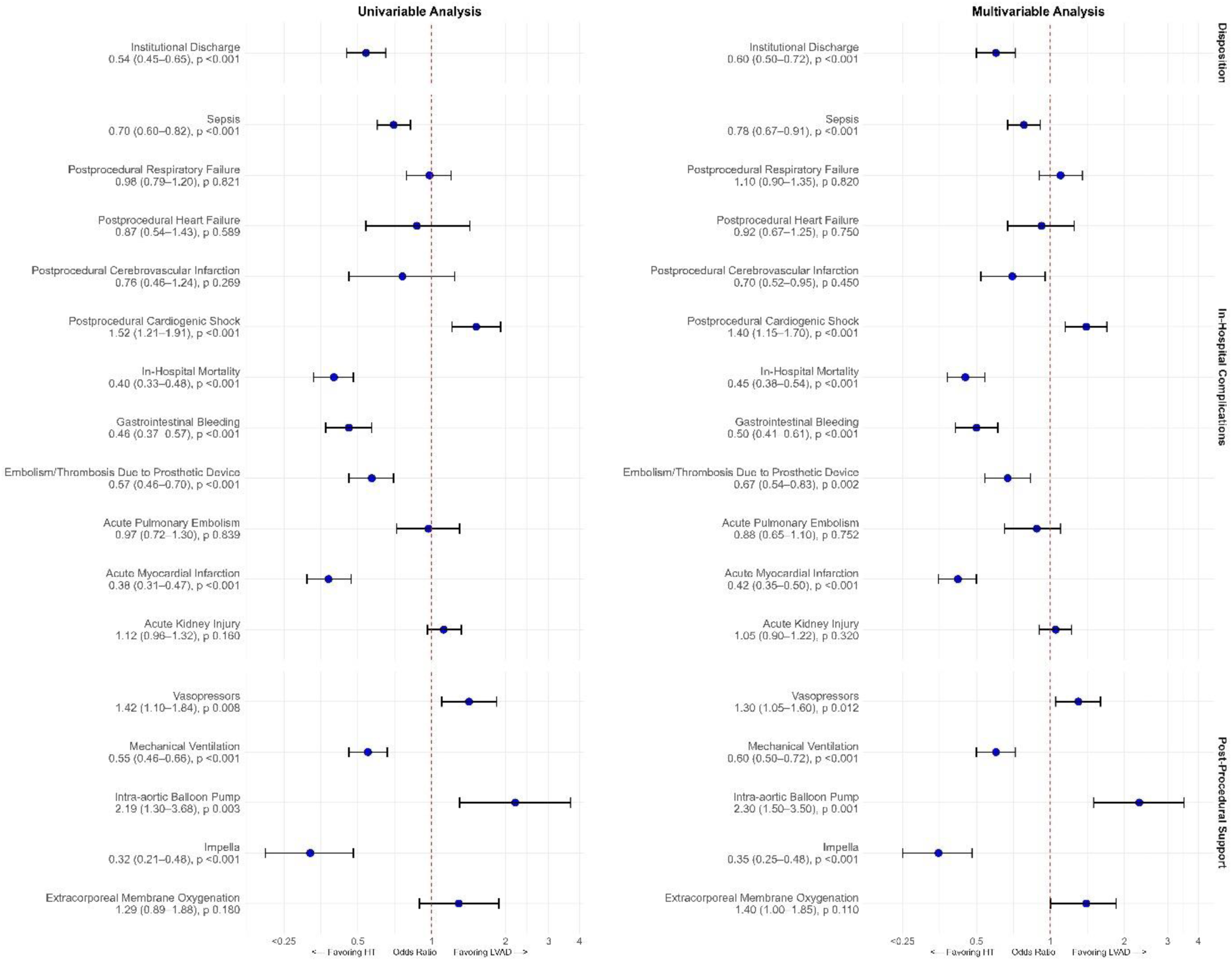
Unadjusted and Adjusted odds of complications amongst HT vs. LVAD during Index hospitalization.

Additionally, patients undergoing HT had a higher need for post-procedure vasopressor support (4.9% vs 3.5%; OR 1.30, 95% CI 1.05 to 1.60, p = 0.012) and post-procedure intra-aortic balloon pump (1.3% vs 0.6%; OR 2.30, 95% CI 1.50 to 3.50, p = 0.001). HT recipients also had higher odds of post-procedure cardiogenic shock (8.7% vs 5.9%; OR 1.40, 95% CI 1.15 to 1.70, p <0.001). However, they required less post-procedure mechanical ventilation (5.3% vs 9.2%; OR 0.60, 95% CI 0.50 to 0.72, p <0.001) and post-procedure Impella support (0.7% vs 2.1%; OR 0.35, 95% CI 0.25 to 0.48, p <0.001).

Patients receiving HT were more likely to have a home discharge (33.5% vs 19.5%, p <0.001) and had lower odds of institutional discharge to a skilled nursing facility (10.8% vs 18.3%, OR 0.60, 95% CI 0.50 to 0.72, p <0.001).

### Financial burden

Annual trends in median index admission costs are shown in **Supplement Figure 6.** Receipt of LVAD bore a higher index hospitalization cost ($237,443 vs $215,350), a difference of $22,093 (p < 0.001) (**Table 1**). Male and female LVAD recipients had median expenses of around $239,215 and $232,240, respectively, compared to their HT counterparts at $218,754 and $207,577. These differences of $20,461 and $24,663, respectively, were lower in HT in both genders (p < 0.001). The cost of index hospitalizations proportionately increased with income quartiles. For LVAD patients with the lowest income quartile, the median index admission cost was $229,898, with the highest quartile seeing a median price of $270,011. This increase was mirrored within the HT patient data as well, where the lowest income quartile incurred a median cost of $196,147, while their highest quartile counterparts faced a median cost of $246,445 **(Supplement Table 2**).

### Propensity matched Analysis

In the secondary analysis using propensity-score matching, including 10,660 patients (5,330 had HT and 5,330 underwent LVAD implantation), receiving HT remained associated with significantly lower in-hospital mortality (5.8% vs 11.9%; p < 0.001), acute gastrointestinal bleeding (3.4% vs 6.9%; p < 0.001), thromboembolic complications due to prosthetic devices (4.0% vs 6.8%; p < 0.001), acute myocardial infarction (4.8% vs 10.0%; p < 0.001), and sepsis (13.5% vs 17.9%; p < 0.001).

Like unmatched analysis, patients receiving HT had significantly higher rates of post-procedural cardiogenic shock (8.9% vs 5.6%; p < 0.001), post-procedure vasopressor use (4.8% vs 3.3%; p = 0.002), ECMO support (3.0% vs 1.7%; p < 0.001), and IABP use (1.6% vs 0.7%; p < 0.001). Moreover, patients who underwent HT required less post-procedure mechanical ventilation (5.8% vs 9.4%; p < 0.001) and Impella support (0.7% vs 2.5%; p < 0.001). Patients receiving HT had a significantly higher likelihood of home discharge (31.9% vs 20.5%; p < 0.001) and a lower probability of discharge to skilled nursing facilities (10.8% vs 16.3%; p < 0.001).

Similarly, resource utilization measures, such as length of stay (median 32 days vs 34 days; p < 0.001), index hospitalization costs (median $228,869 vs $246,082; p < 0.001), and post-procedural length of stay (17 vs 21 days; p < 0.001), were significantly lower in HT compared to LVAD recipients.

### Readmission Characteristics

The temporal distribution until the first readmission for HT and LVAD recipients, 30-day and 90-day first, and cumulative readmission rates are shown in **Central Illustration**.

### 30-day readmission characteristics

We identified 22,753 weighted HT and LVAD recipients who survived discharge from the index hospitalizations; 11,409 (50.1%) had LVAD. A higher proportion of LVAD recipients were re-hospitalized within 30 days (27.9% vs. 25%, p=0.02), and had a longer LOS (6 vs 5 days, p < 0 .001) compared with HT recipients (**Table 2**). As compared to patients who did not sustain 30-day readmissions, patients who required 30-day readmissions were older, had higher costs and longer LOS during index hospitalizations, had Medicare, diabetes with complications, prior thromboembolic events, and history of malignancy in both LVAD and HT groups. (**Supplement Table 5**).

**Table 2:** 30-day and 90-day readmission characteristics and In-Hospital Outcomes of HT and LVAD recipients.

|  |  |  |  |  |  |  |
| --- | --- | --- | --- | --- | --- | --- |
| Total population January – November<br>Total, n | 0-30-day First readmission number, n | 0–30-day First readmission rate | Length of stay (LOS), median, IQR | Procedures, median, IQR | Total cost per readmission |  |
| HT*, n = 11344 | 2836 | 25% | 5 (3–9) | 2 (1–4) | \$16,606 (\$9,471–\$31,144) | |
| LVAD*, n = 11409 | 3182 | 27.9% | 6 (3–11) | 1 (0–3) | \$11,736 (\$6,017–\$27,067) | |
| p-value |  | 0.02 | < 0 .001 | < 0 .001 | < 0 .001 |  |
| Total population January – September<br>Total, n | 0-90-day First readmission number, n | 0–90-day First readmission rate | Length of stay, median (LOS), IQR | Procedures, median, IQR | Total cost per readmission |  |
| HT*, n = 9341 | 3671 | 39.3% | 5 (2–8) | 2 (1–4) | \$15,233 (\$8,400–\$28,581) | |
| LVAD*, n = 9283 | 4172 | 45% | 6 (3–11) | 1 (0–3) | \$11,680 (\$6,111–\$26,163) | |
| p-value |  | < 0 .001 | < 0 .001 | < 0 .001 | < 0 .001 |  |
| Propensity Matched<br>Total population January – November<br>Total, n | 0-30-day Propensity-matched readmission number, n | 0–30-day Propensity-matched readmission rate | Length of stay (LOS), median, IQR | Procedures, median, IQR | Total cost per readmission |  |
| HT*, n = 4136 | 1062 | 25.6% | 5 (3 – 9) | 2 (1-5) | \$ 17541 (\$ 9613 - \$ 34575) | |
| LVAD*, n = 4136 | 1158 | 27.9% | 5 (3 – 11) | 1 (0-3) | \$ 12731 (\$ 6424 - \$ 28983) | |
| p-value |  | 0.09 | < 0 .001 | < 0 .001 | < 0 .001 |  |
| Propensity Matched<br>Total population January – September<br>Total, n | 0-90-day Propensity-matched readmission number, n | 0–90-day Propensity-matched readmission rate | Length of stay (LOS), median, IQR | Procedures, median, IQR | Total cost per readmission |  |
| HT*, n = 3392 | 1381 | 40.7% | 5 (2–8) | 2 (1–4) | \$16,237 (\$8,833–\$29,438) | |
| LVAD*, n = 3392 | 1549 | 45.6% | 6 (3–11) | 1 (0–3) | \$12,635 (\$6,354–\$28,166) | |
| p-value |  | 0.004 | < 0 .001 | < 0 .001 | < 0 .001 |  |
| In-hospital outcomes amongst HT and LVAD during Index hospitalization |  |  |  |  |  |  |
|  | Unmatched Cohort |  |  | Propensity matched |  |  |
|  | HT | LVAD | P – value | HT | LVAD | P - value |
| Resource utilization, Median (IQR) |  |  |  |  |  |  |
| LOS (days) | 31 (17 – 53) | 33 (23 – 50) | < 0.001 | 32 (18–56) | 34 (23–51) | < 0.001 |
| Index Hospitalization Costs in US Dollars | \$ 215350 (\$ 146340 - \$ 335018) | \$ 237443 (\$ 178912 - \$ 334167) | < 0.001 | \$ 228869 (\$ 154118 – \$ 363122) | \$ 246082 (\$ 185086 - \$ 354230) | < 0.001 |
| Post-procedural LOS | 17 (12 – 27) | 21 (14 – 34) | <0.001 | 17 (12 – 28) | 21 (15 – 35) | <0.001 |
| Procedures on Index Admission | 13 (7 – 20) | 12 (6 – 19) | < 0.001 | 13 (7–21) | 11 (6–18) | <0.001 |
| Complications |  |  |  |  |  |  |
| In-Hospital Mortality | 718 (5.5%) | 1818 (12.71%) | < 0.001 | 308 (5.78%) | 635 (11.91%) | <0.001 |
| Embolism/thrombosis Due to Prosthetic device | 533 (4.10%) | 1,000 (6.97%) | < 0.001 | 213 (3.99%) | 364 (6.83%) | <0.001 |
| Acute pulmonary embolism | 307 (2.36%) | 348 (2.43%) | 0.83 | 127 (2.38%) | 142 (2.66%) | 0.46 |
| Sepsis | 1,707 (13.12%) | 2,533 (17.72%) | < 0.001 | 719 (13.49%) | 956 (17.94%) | <0.001 |
| Post-procedural respiratory failure | 1,188 (9.13%) | 1,334 (9.33%) | 0.82 | 488 (9.16%) | 465 (8.72%) | 0.60 |
| Post-procedural Heart Failure | 130 (1.00%) | 163 (1.14%) | 0.59 | 47 (0.88%) | 53 (0.99%) | 0.61 |
| Post-procedural Cerebrovascular infarction | 72 (0.55%) | 104 (0.73%) | 0.27 | 33 (0.62%) | 50 (0.94%) | 0.13 |
| Gastrointestinal Bleeding | 432 (3.32%) | 995 (6.96%) | < 0.001 | 181 (3.40%) | 366 (6.87%) | <0.001 |
| Acute Myocardial Infarction | 555 (4.27%) | 1,500 (10.50%) | < 0.001 | 255 (4.78%) | 534 (10.02%) | <0.001 |
| Acute Kidney Injury | 9,930 (76.31%) | 10,602 (74.16%) | 0.16 | 4,130 (77.49%) | 3895 (73.08%) | 0.002 |
| Post-procedural cardiogenic shock | 1,136 (8.73%) | 844 (5.90%) | <0.001 | 472 (8.86%) | 300 (5.63%) | <0.001 |
| **Discharge Disposition** |  |  |  |  |  |  |
| Institutional Discharge | 1,411(10.8%) | 2,623(18.3%) | < 0.001 | 575 (10.79%) | 870 (16.32%) | < 0.001 |
| Home discharge | 4,363(33.5%) | 2,794(19.5%) | < 0.001 | 1700 (31.89%) | 1094 (20.53%) | < 0.001 |
|  | HT | LVAD |  | HT | LVAD |  |
| **Hemodynamic/Respiratory support** | Unmatched |  |  | Propensity matched |  |  |
| Post-procedure vasopressors | 641 (4.9%) | 503 (3.5%) | 0.007 | 254 (4.8%) | 174 (3.3%) | 0.002 |
| Post-procedure ECMO | 349 (2.7%) | 298 (2.1%) | 0.17 | 162 (3.0%) | 93 (1.7%) | < 0.001 |
| Post-procedure Impella | 87 (0.7%) | 297 (2.1%) | < 0.001 | 39 (0.7%) | 133 (2.5%) | < 0.001 |
| Post-procedure IABP | 90 (0.6%) | 178 (1.3%) | 0.002 | 87 (1.6%) | 39 (0.7%) | < 0.001 |
| Post-procedure Mechanical ventilation | 687 (5.3%) | 1,318 (9.2%) | < 0.001 | 311 (5.8%) | 502 (9.4%) | < 0.001 |

\*Discharged alive after index hospitalization. LOS: Length of Stay; ECMO: Extracorporeal Membrane Oxygenation; IABP: Intra-aortic Balloon Pump

**Figure 3 (Panel A & C)** presents the relative frequency of primary diagnoses associated with 30-day readmissions. Heart failure (24.1%), device complications (13.7%), gastrointestinal bleeding (9.9%), and other causes (43.6%) accounted for most readmissions in LVAD recipients. In comparison, complications of transplant (29.8%), renal disorder (6.2%), and other causes (42.7%) accounted for most readmissions amongst HT recipients. Importantly, the rate of 30-day readmission varied from 6% to 39%, and 0%-40%, by hospital deciles in readmission amongst HT and LVAD recipients, respectively (**Supplement Figure 7**).

**Figure 3:**
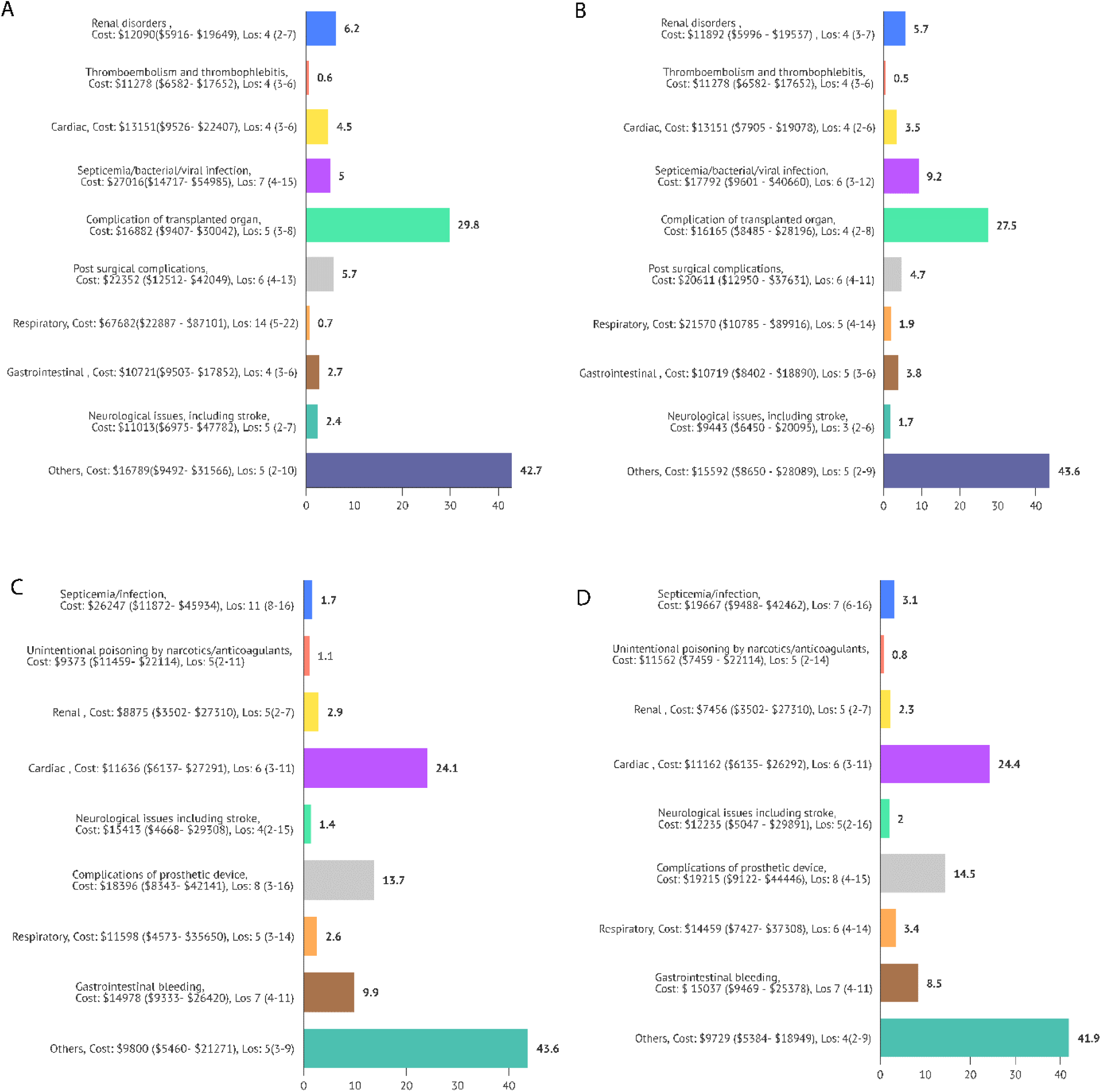
Reasons, Hospital Costs, and Length of Stay for 30-day First Readmission in HT Recipients (A) and LVAD Recipients (C). Reasons, Hospital Costs, and Length of Stay for 90-day First Readmission into HT Recipients (B) and LVAD Recipients (D)

### Financial burden

HT recipients had a higher median readmission cost [$16,606 ($9,471 - $31,144) vs $11,736 ($6,017 - $27,067)], a difference of $4,870 favoring LVAD (p < 0.001). This may correlate with differences in the number of procedures (2 vs 1, p <0.001) (**Table 2**). Specifically, male HT recipients ($16,942; $9,526 – $30,042) incurred higher median readmission costs than their LVAD counterparts ($11,872; $6,135 - $27,291) with a difference of $5,070; p<0.001). Furthermore, Medicare payments were $4,918 higher for HT ($16,967, $9,919 – $33,688) than LVAD ($12,049, $5,996 - $27,156; p< 0.001). We noticed a similar increase in readmission costs among income quartiles. Individuals in the 76th – 100th percentile of median household income exhibited the highest readmission costs for both procedures when compared to patients in the lower income quartile group. (**Supplement Table 3**).

Among HT recipients, 30-day readmission increased the cumulative cost by 18% (β coefficient, 0.18; 95% confidence interval, 0.16, 0.21; p < 0.001). Among other covariates, utilization of ECMO, Impella, IABP and mechanical ventilation, and co-morbidities including hypertension, chronic liver disease are associated with higher costs. Similarly, in LVAD recipients, 30-day readmission increased the cumulative cost by 15% (β coefficient, 0.15; 95% confidence interval, 0.12, 0.17; p < 0.001). Among other covariates, utilization of ECMO, Impella, IABP, mechanical ventilation, and co-morbidities including hypertension, and chronic liver disease are associated with higher costs (**Table 3 & 4)**.

**Table 3:**
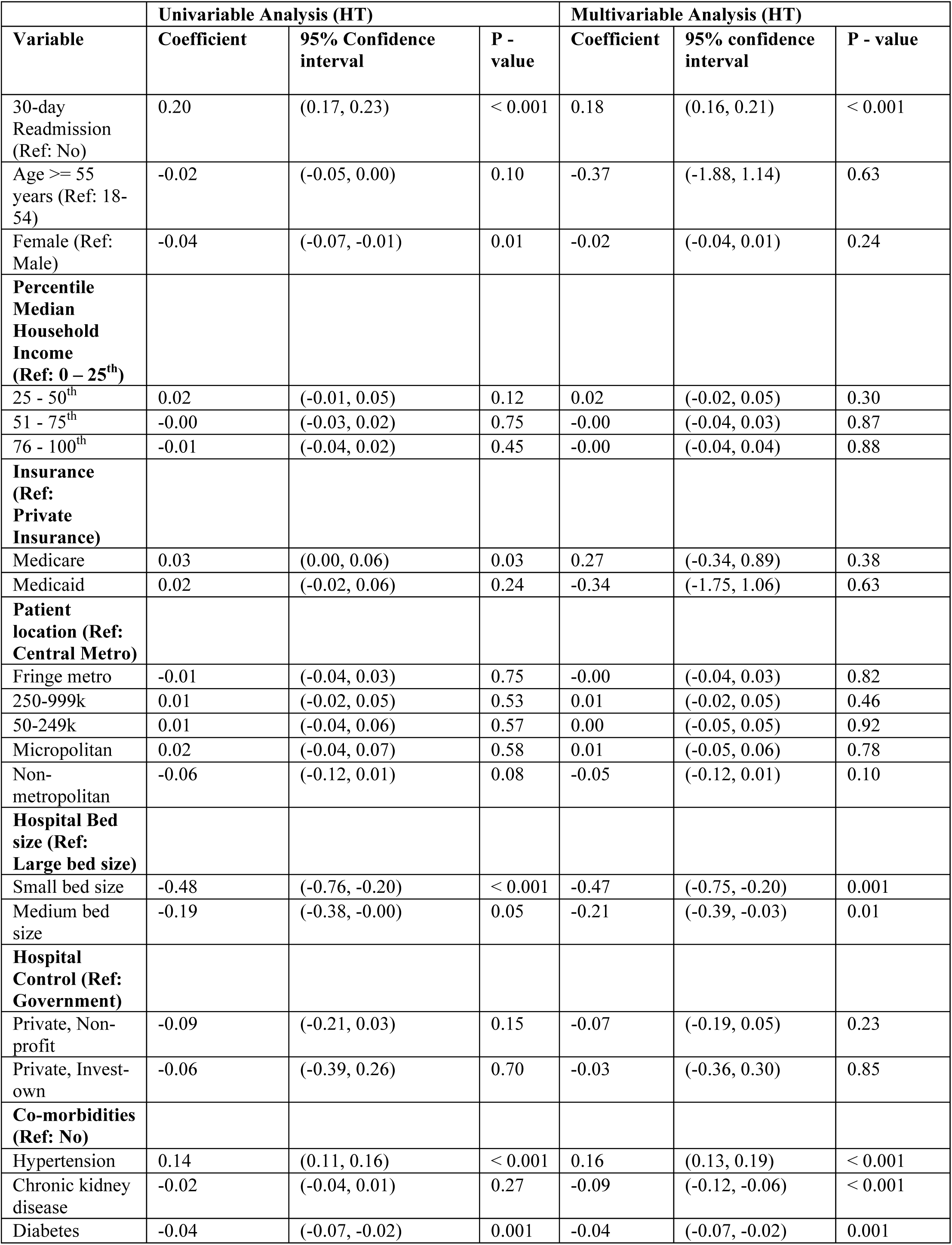
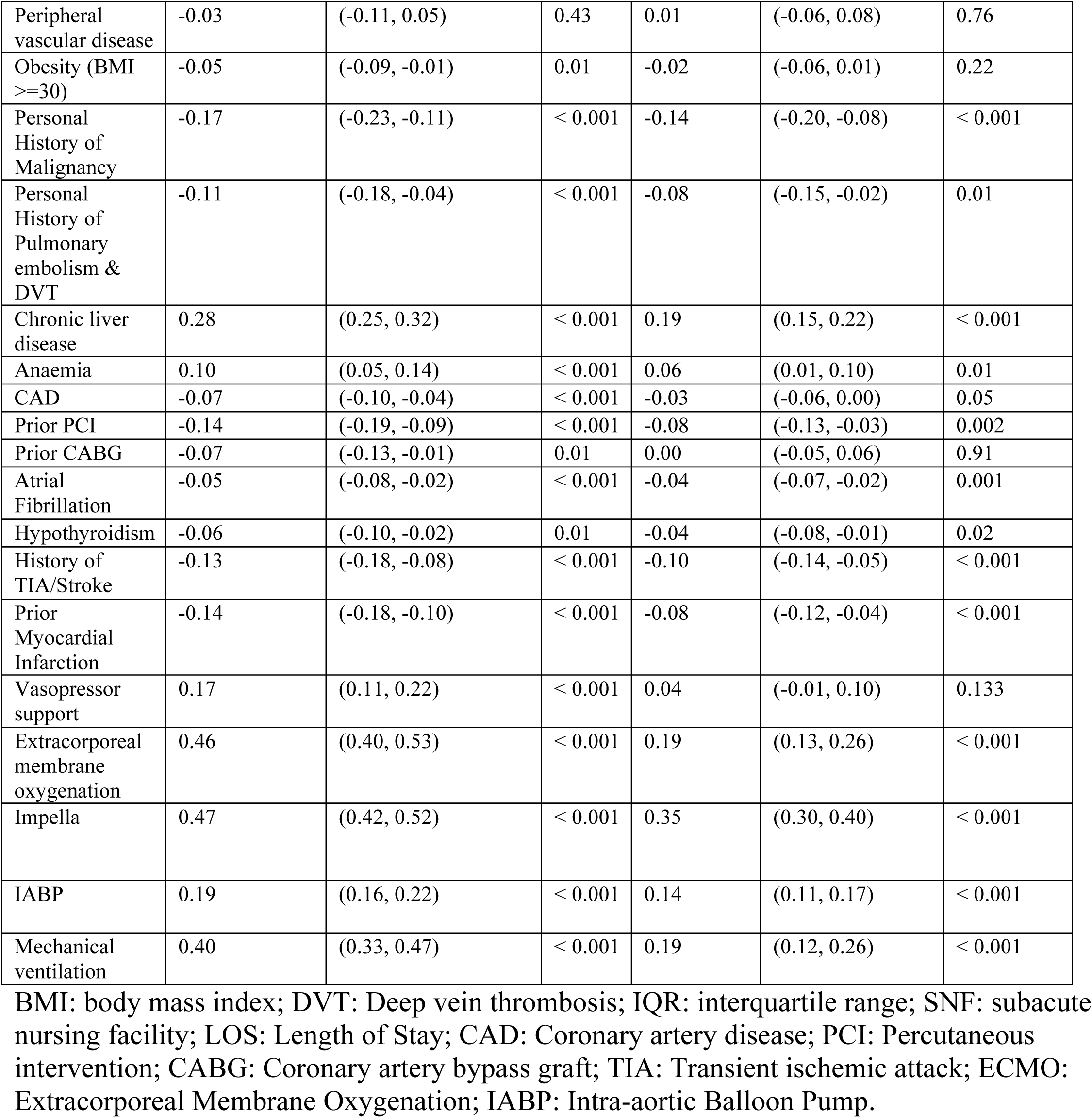
Univariable and Multivariable logistic Regression Analysis to Assess the Association of Readmission within 30 days on total cost of heart transplantation.

|  | Univariable Analysis (HT) |  |  | Multivariable Analysis (HT) |  |  |
| --- | --- | --- | --- | --- | --- | --- |
| Variable | Coefficient | 95% Confidence interval | P - value | Coefficient | 95% confidence interval | P - value |
| 30-day Readmission (Ref: No) | 0.20 | (0.17, 0.23) | < 0.001 | 0.18 | (0.16, 0.21) | < 0.001 |
| Age >= 55 years (Ref: 18-54) | -0.02 | (-0.05, 0.00) | 0.10 | -0.37 | (-1.88, 1.14) | 0.63 |
| Female (Ref: Male) | -0.04 | (-0.07, -0.01) | 0.01 | -0.02 | (-0.04, 0.01) | 0.24 |
| **Percentile Median Household Income (Ref: 0 – 25<sup>th</sup>)** |  |  |  |  |  |  |
| 25 - 50<sup>th</sup> | 0.02 | (-0.01, 0.05) | 0.12 | 0.02 | (-0.02, 0.05) | 0.30 |
| 51 - 75<sup>th</sup> | -0.00 | (-0.03, 0.02) | 0.75 | -0.00 | (-0.04, 0.03) | 0.87 |
| 76 - 100<sup>th</sup> | -0.01 | (-0.04, 0.02) | 0.45 | -0.00 | (-0.04, 0.04) | 0.88 |
| **Insurance (Ref: Private Insurance)** |  |  |  |  |  |  |
| Medicare | 0.03 | (0.00, 0.06) | 0.03 | 0.27 | (-0.34, 0.89) | 0.38 |
| Medicaid | 0.02 | (-0.02, 0.06) | 0.24 | -0.34 | (-1.75, 1.06) | 0.63 |
| **Patient location (Ref: Central Metro)** |  |  |  |  |  |  |
| Fringe metro | -0.01 | (-0.04, 0.03) | 0.75 | -0.00 | (-0.04, 0.03) | 0.82 |
| 250-999k | 0.01 | (-0.02, 0.05) | 0.53 | 0.01 | (-0.02, 0.05) | 0.46 |
| 50-249k | 0.01 | (-0.04, 0.06) | 0.57 | 0.00 | (-0.05, 0.05) | 0.92 |
| Micropolitan | 0.02 | (-0.04, 0.07) | 0.58 | 0.01 | (-0.05, 0.06) | 0.78 |
| Non-metropolitan | -0.06 | (-0.12, 0.01) | 0.08 | -0.05 | (-0.12, 0.01) | 0.10 |
| **Hospital Bed size (Ref: Large bed size)** |  |  |  |  |  |  |
| Small bed size | -0.48 | (-0.76, -0.20) | < 0.001 | -0.47 | (-0.75, -0.20) | 0.001 |
| Medium bed size | -0.19 | (-0.38, -0.00) | 0.05 | -0.21 | (-0.39, -0.03) | 0.01 |
| **Hospital Control (Ref: Government)** |  |  |  |  |  |  |
| Private, Non-profit | -0.09 | (-0.21, 0.03) | 0.15 | -0.07 | (-0.19, 0.05) | 0.23 |
| Private, Invest-own | -0.06 | (-0.39, 0.26) | 0.70 | -0.03 | (-0.36, 0.30) | 0.85 |
| **Co-morbidities (Ref: No)** |  |  |  |  |  |  |
| Hypertension | 0.14 | (0.11, 0.16) | < 0.001 | 0.16 | (0.13, 0.19) | < 0.001 |
| Chronic kidney disease | -0.02 | (-0.04, 0.01) | 0.27 | -0.09 | (-0.12, -0.06) | < 0.001 |
| Diabetes | -0.04 | (-0.07, -0.02) | 0.001 | -0.04 | (-0.07, -0.02) | 0.001 |
| Peripheral vascular disease | -0.03 | (-0.11, 0.05) | 0.43 | 0.01 | (-0.06, 0.08) | 0.76 |
| Obesity (BMI $\geq 30$) | -0.05 | (-0.09, -0.01) | 0.01 | -0.02 | (-0.06, 0.01) | 0.22 |
| Personal History of Malignancy | -0.17 | (-0.23, -0.11) | < 0.001 | -0.14 | (-0.20, -0.08) | < 0.001 |
| Personal History of Pulmonary embolism & DVT | -0.11 | (-0.18, -0.04) | < 0.001 | -0.08 | (-0.15, -0.02) | 0.01 |
| Chronic liver disease | 0.28 | (0.25, 0.32) | < 0.001 | 0.19 | (0.15, 0.22) | < 0.001 |
| Anaemia | 0.10 | (0.05, 0.14) | < 0.001 | 0.06 | (0.01, 0.10) | 0.01 |
| CAD | -0.07 | (-0.10, -0.04) | < 0.001 | -0.03 | (-0.06, 0.00) | 0.05 |
| Prior PCI | -0.14 | (-0.19, -0.09) | < 0.001 | -0.08 | (-0.13, -0.03) | 0.002 |
| Prior CABG | -0.07 | (-0.13, -0.01) | 0.01 | 0.00 | (-0.05, 0.06) | 0.91 |
| Atrial Fibrillation | -0.05 | (-0.08, -0.02) | < 0.001 | -0.04 | (-0.07, -0.02) | 0.001 |
| Hypothyroidism | -0.06 | (-0.10, -0.02) | 0.01 | -0.04 | (-0.08, -0.01) | 0.02 |
| History of TIA/Stroke | -0.13 | (-0.18, -0.08) | < 0.001 | -0.10 | (-0.14, -0.05) | < 0.001 |
| Prior Myocardial Infarction | -0.14 | (-0.18, -0.10) | < 0.001 | -0.08 | (-0.12, -0.04) | < 0.001 |
| Vasopressor support | 0.17 | (0.11, 0.22) | < 0.001 | 0.04 | (-0.01, 0.10) | 0.133 |
| Extracorporeal membrane oxygenation | 0.46 | (0.40, 0.53) | < 0.001 | 0.19 | (0.13, 0.26) | < 0.001 |
| Impella | 0.47 | (0.42, 0.52) | < 0.001 | 0.35 | (0.30, 0.40) | < 0.001 |
| IABP | 0.19 | (0.16, 0.22) | < 0.001 | 0.14 | (0.11, 0.17) | < 0.001 |
| Mechanical ventilation | 0.40 | (0.33, 0.47) | < 0.001 | 0.19 | (0.12, 0.26) | < 0.001 |

BMI: body mass index; DVT: Deep vein thrombosis; IQR: interquartile range; SNF: subacute nursing facility; LOS: Length of Stay; CAD: Coronary artery disease; PCI: Percutaneous intervention; CABG: Coronary artery bypass graft; TIA: Transient ischemic attack; ECMO: Extracorporeal Membrane Oxygenation; IABP: Intra-aortic Balloon Pump.

**Table 4:**
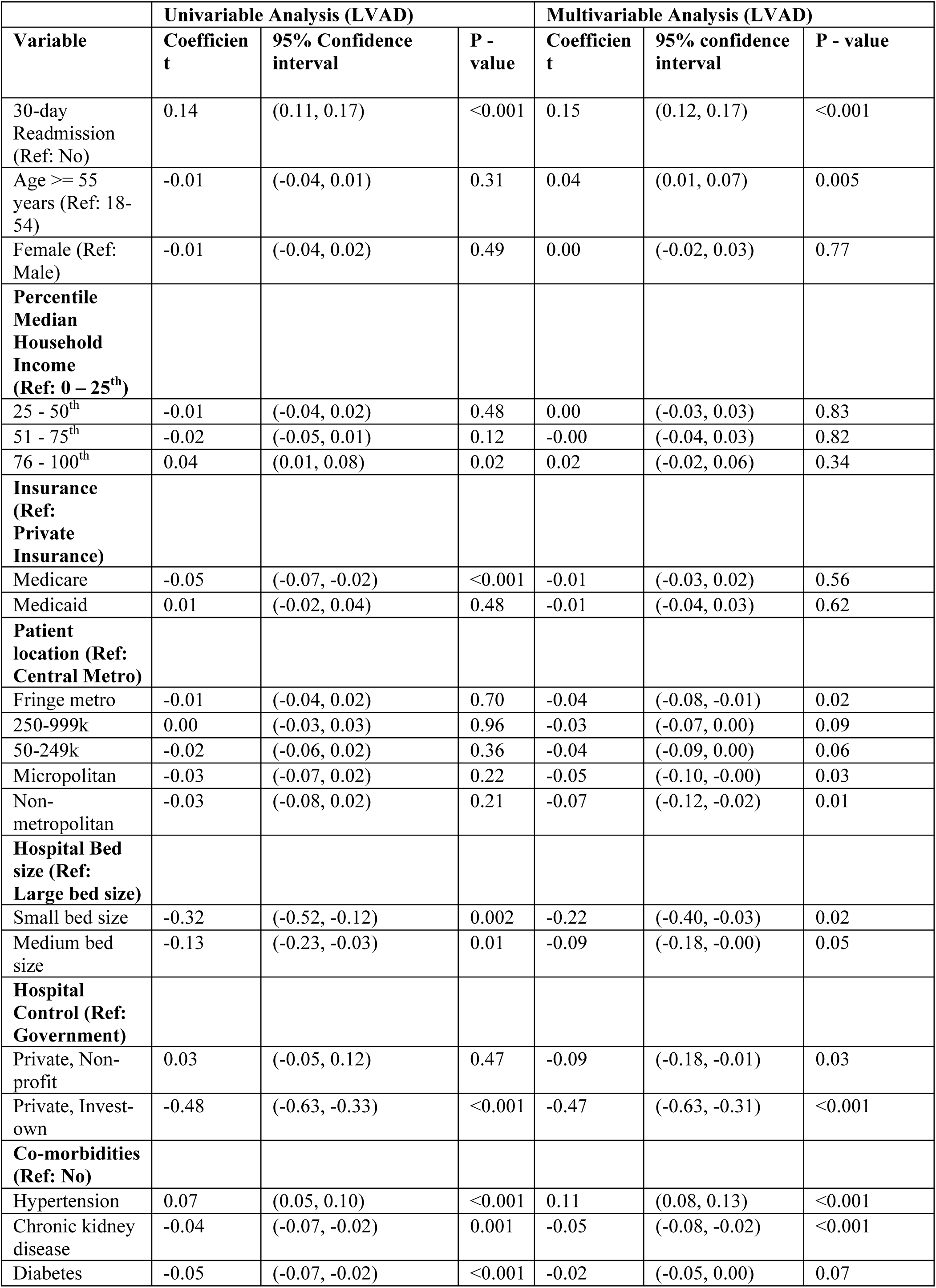
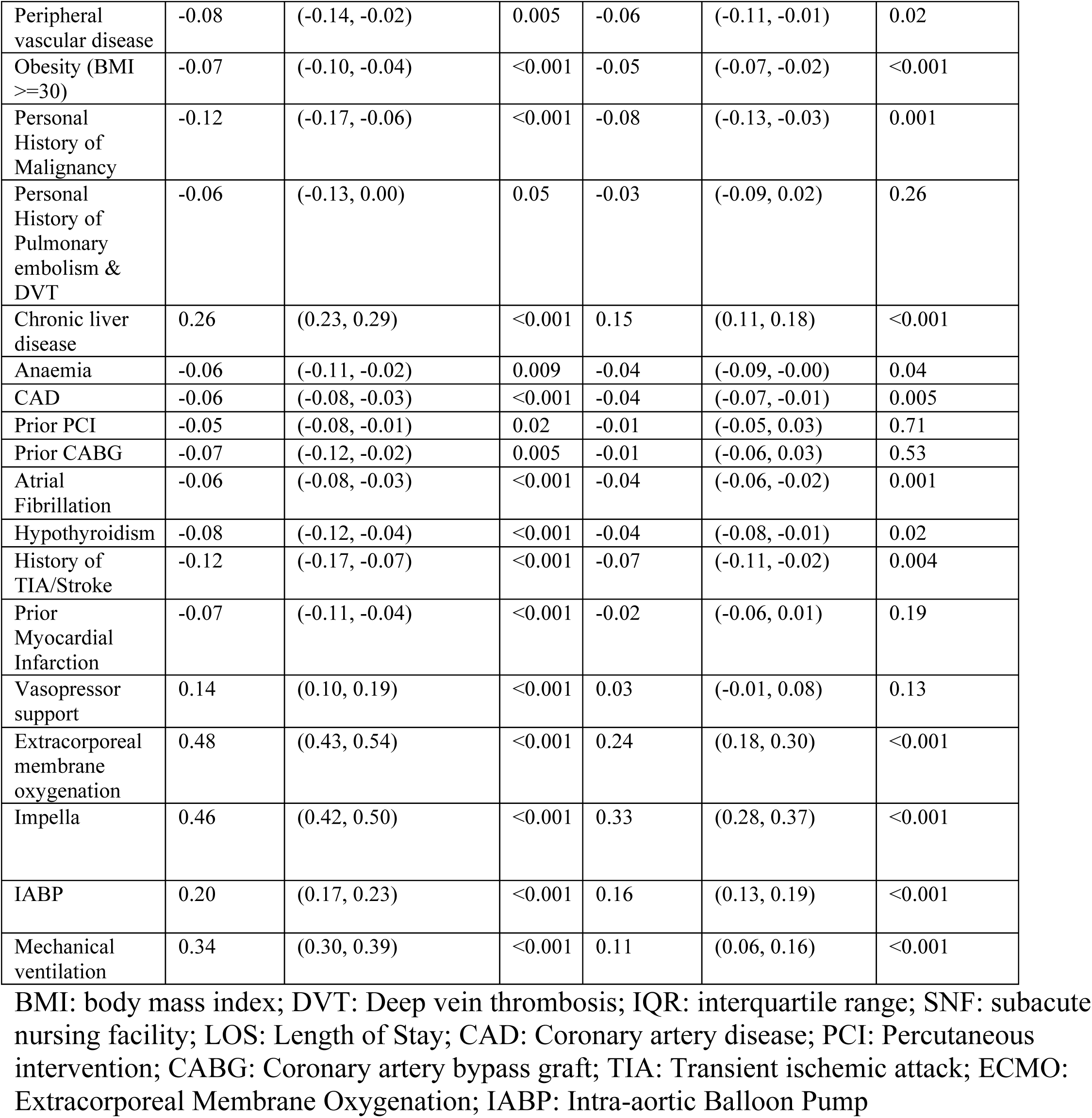
Univariable and Multivariable Logistic Regression Analysis to Assess the Association of Readmission within 30 days on total cost of LVAD.

We performed a sensitivity analysis to assess the differences in costs for index HT and LVAD hospitalizations among patients with and without 30-day readmissions. Multivariable hierarchical logistic regression analysis revealed that 30-day readmission in the HT cohort was associated with an 8% increase in costs (p<0.001, 95% CI: 5% to 10%). Among covariates, utilization of ECMO, Impella, IABP, and mechanical ventilation, as well as chronic medical conditions including hypertension, chronic liver disease, and anemia, were associated with higher costs. In comparison, 30-day readmission in the LVAD cohort was associated with a 5% increase in costs (p<0.001, 95% CI: 2% to 7%). Among other covariates, age ≥ 55 years, utilization of ECMO, Impella, IABP, and mechanical ventilation, as well as chronic medical conditions including hypertension and chronic liver disease, were associated with higher costs (**Table 5**).

**Table 5:**
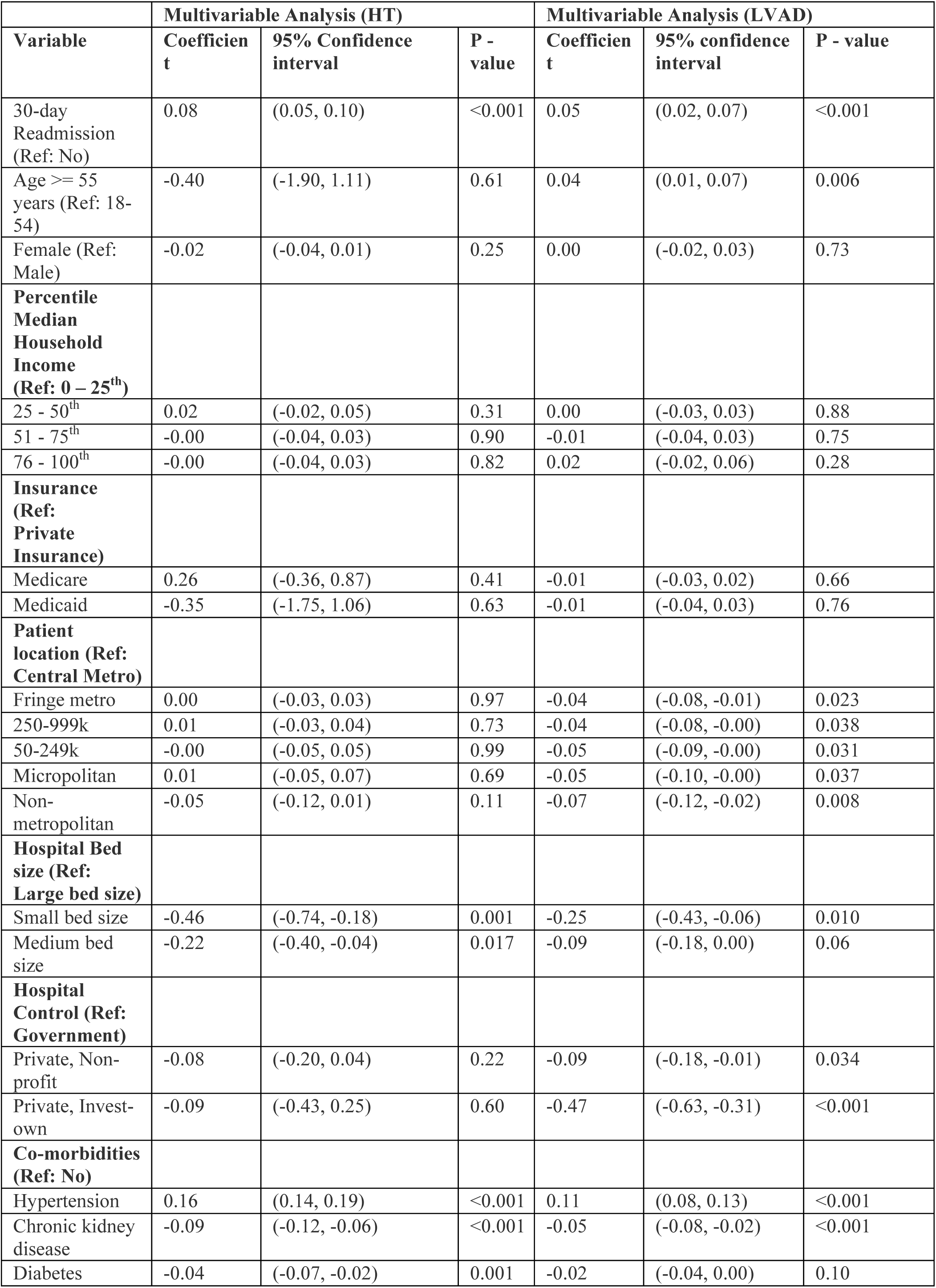
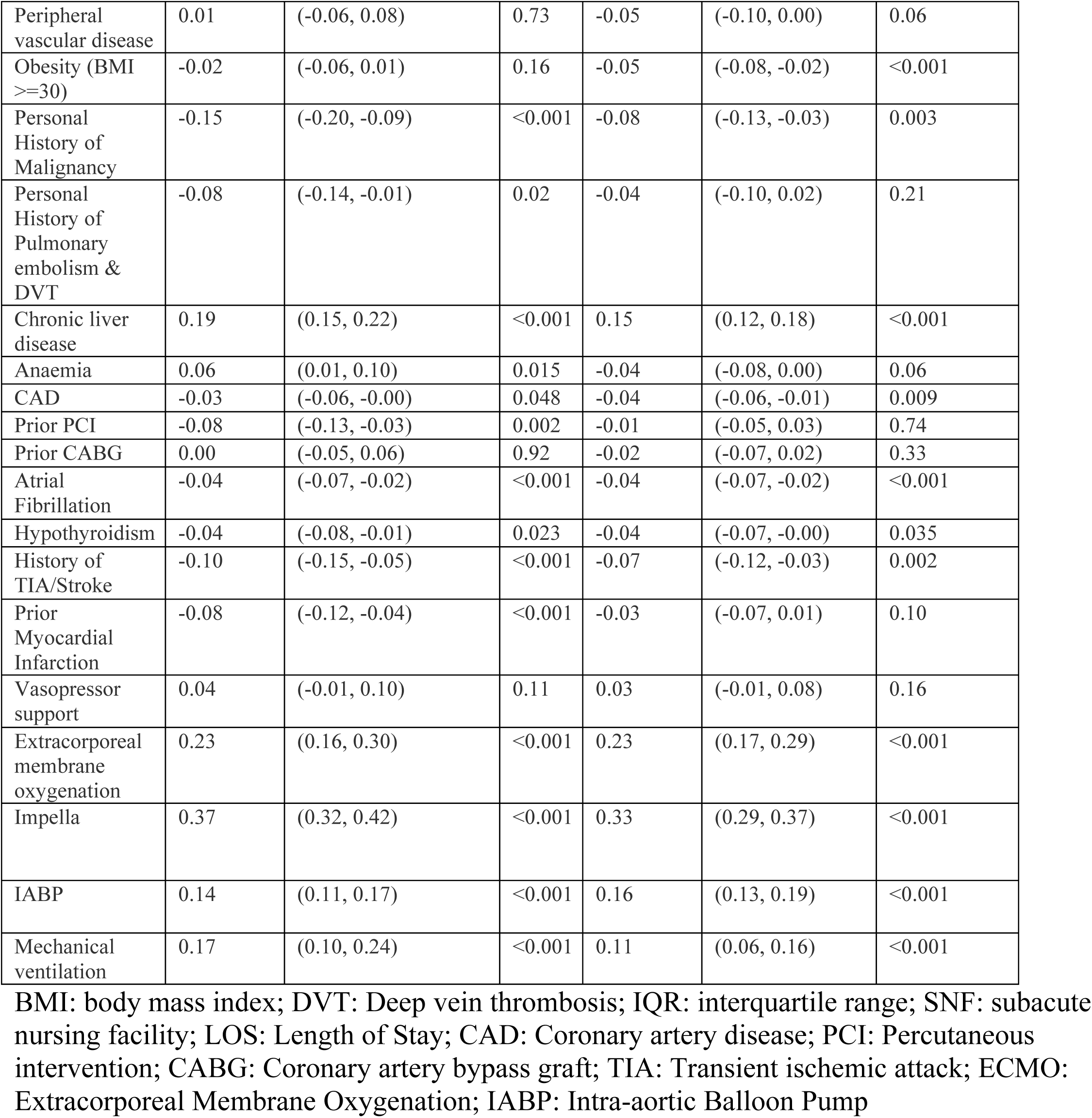
Multivariable hierarchal logistic regression models to assess the difference in index HT and LVAD costs between those with 30-day readmission versus no readmission.

### 90-day readmission characteristics

We identified 18,624 weighted HT and LVAD recipients included in 90-DRC; 9283 (49.8%) had LVADs. Among these, LVAD recipients were significantly more likely to be hospitalized within 90 days (4172, 45% vs. 3671, 39.3%; p < 0.001). LVAD recipients had a longer hospital stay (5 vs 6 days, p = 0.001). However, HT recipients underwent more procedures during readmissions (2 vs 1, p <0.001) (**Table 2**).

Compared to patients who did not sustain 90-day readmissions, patients who required 90-day readmissions were older, female, had higher costs, longer LOS and had undergone more procedures during index hospitalizations, had Medicare, had diabetes with complications, prior thromboembolic events, and history of malignancy in both LVAD and HT groups (**Supplement Table 6**).

**Figure 3 (Panel B&D)** presents the relative frequency of primary diagnoses associated with 30-day readmissions. Heart failure (24.4%), device complications (14.5%), gastrointestinal bleeding (8.5%), and other causes (41.9%) accounted for most readmissions in LVAD recipients. In comparison, complications of transplant (27.5%), sepsis (9.2%), renal disorder (5.7%), and other causes (43.6%) accounted for most readmissions amongst HT recipients. Importantly, the rate of 90-day readmission varied from 8% to 46%, and 2%-49%, by hospital deciles in readmission amongst HT and LVAD recipients, respectively (**Supplement Figure 7**).

Recipients of HT had a higher median readmission cost [$15,233 ($8,400–$28,581) vs. $11,680 ($6,111–$26,163)] which is a difference of $3,551 favoring LVAD (p=0.001) (**Table 2**).

### Predictors for readmission and costs

On multivariable hierarchical logistic regression analysis female sex, and longer index hospital stays (≥30 days) were associated with higher odds of 30 & 90-day readmissions among HT recipients. Among LVAD recipients, longer index hospital stays (≥30 days), Medicare insurance, peripheral vascular disease, pre-procedural vasopressor support, and use of IABP were associated with higher odds of 30 & 90-day readmissions **(Supplement Table 7)**.

## Discussion

Patients seek treatment with HT and LVADs to mitigate the symptoms, morbidity, and mortality of refractory HF. Survival after such therapies constitutes part of the performance evaluation by the Centers of Medicare and Medicaid Services as well as private insurances, and hence are widely discussed with the patients. However, the absence of reporting for hospitalizations, care costs, and non-fatal complications following these therapies results in unintended consequences: lack of data around these outcomes beyond clinical trials, which are limited, and a lack of informed discussion between patients and providers.

In this contemporary analysis, we have identified characteristics of HT and LVAD recipients, complications following HT or LVAD implantation, economic burden, re-hospitalization risks and predictors in a nationally representative NRD cohort in the United States. Our results are important given that there is limited real-world data to inform patients and providers about short-term expectations from selected forms of therapy, beyond long term survival.

Our major findings indicate both LVAD and HT recipients sustain significant readmissions and associated costs of care. In the United States, LVAD recipients had higher index hospitalization costs ($22,115 higher), lengths of stay (2 days longer), and readmission rates when compared to HT recipients in the United States. Moreover, as compared to HT recipients, LVAD recipients were more likely to come from lower-income groups and have Medicare insurance. Other findings included: the increasing cost of index hospitalization with rising recipient income for both LVAD and HT recipients; the lower rehospitalization costs among LVAD recipients; and the identification of LVAD and HT-related complications that accounted for the significant risk of rehospitalizations **(Central Illustration).** In addition, our results also highlight a wide variation in readmission rates between the highest and lowest hospital deciles.

Inherently, HT and LVAD therapies are costly interventions and hence would incur high costs of index hospitalization, in addition to the cost of hospitalization while candidacy is being considered, and acute illness is being addressed. Beyond this, our observation of high index hospitalization mortality (12.7%), acute myocardial infarction (10%), sepsis (17%), post-procedure respiratory failure (9%) and GI bleeding and thromboembolic events (each 7%) amongst LVAD recipients and high respiratory failure (9%), sepsis (>13%) and post-operative cardiogenic shock (9%) in HT recipients point to potential opportunities to identify high-risk patients to reduce morbidity and cost during index hospitalization for both HT and LVAD. More than half of the LVAD and HT recipients in the US sustained hospital readmissions at 30- and 90-days, LVAD recipients with an even higher trend. Moreover, the mean cumulative costs for those who were readmitted were around 20% higher than those who did not have a readmission in both HT and LVAD recipients. This suggests a substantial financial impact of readmissions after HT and LVAD; however, it is important to review these results in the context of whether these readmissions could be attributed to HT and LVAD. Gastrointestinal bleeding and residual heart failure contributed to a significant number of readmissions in the LVAD group while HT-specific compilations-related hospitalizations were also likely related to immunosuppression-related AKI, infections, and possibly rejection suspicion/management.

Numerous studies, including our own, have identified gastrointestinal bleeding as an important cause of rehospitalization.^7^Developing ways to mitigate these complications could, therefore, help cut costs significantly. Recently, the antiplatelet removal and hemocompatibility events with the HeartMate 3 pump (ARIES-HM3) trial showed nearly 40% reduction in GI bleeding events without an increase in thrombosis or stroke alongside 47% reduction in hospital days due to bleeding events as well as 41% reduction in cost of care for bleeding events with discontinuation of aspirin in HM3 recipients.^8^Based on these results, the FDA approved HM3 LVAD use without aspirin on August 21, 2024.

The use of direct oral anticoagulants (DOAC) has improved both thrombosis and bleeding complications in atrial fibrillation. However, the use of DOAC has been limited among LVAD recipients until recently. Emerging data is showing promising results supporting not only safety but also effectiveness in LVAD recipients as a substitute for warfarin.^9,10^ As these early findings continue to find validation in randomized controlled trials, there is hope that the use of DOAC has the potential to reduce GI bleeding and associated morbidity which remains a major cause of rehospitalization in LVAD recipients.^11^ Similarly, analysis of the INTERMACS database suggests that elderly patients, who are typically at the highest risk of bleeding, are at a relatively low risk of LVAD thrombosis, suggesting that an individualized antithrombotic regimen might be safe and effective.^12^ Beyond considerations in anticoagulation management, usage of guideline-directed medical therapy (GDMT) may improve outcomes in patients with end-stage heart failure, including LVAD recipients.^13,14^ These measures could result in significantly improved patient outcomes and cost-savings to the system, further making LVAD more comparable to HT.

On the other hand, infection and renal injury-related hospitalizations in HT recipients also present opportunities to reduce the cost and burden of care. Increased risk in HT recipients likely reflects the increased susceptibility to infection and nephrotoxic effect stemming from ongoing immunosuppression adjustments and aiming for higher doses/levels during early periods post-HT.^15^ Studies exploring decreased or rapidly deescalated immunosuppression and better surveillance methods might thus contribute to fewer downstream readmissions.^16^

Our study noted that Medicare and Medicaid are the primary payers for nearly half of the nation’s HT and two-thirds of LVAD hospitalizations, highlighting the pertinence of our findings to public health. The expansion of Medicaid led to a significant increase in rates of LVAD implantation, most prominently among African American men.^17^ However, these coverages might be subject to change with increasing enrollment and when rising costs become prohibitive. Although reducing costs in pursuit of high-value cost-conscious care is essential, reduced funding for these procedures through the CMS may adversely affect millions, particularly those from socio-culturally disadvantaged groups.^18^

We observed differences in the use of HT and LVADs by socioeconomic status (SES) that may, in part, be explained by the influence of SES on clinical outcomes. While lower SES was associated with worse outcomes following HT, it did not significantly impact LVAD outcomes.^19,20^ Individuals from such vulnerable groups, particularly Black individuals, are known to be less likely to receive transplants and have higher risks of waitlist mortality.^21^ Risks of post-transplantation death in this group are also known to be higher.^22^ Conversely, outcomes with LVADs in Black patients have been demonstrably better than other demographics, suggesting that these devices may play a pivotal role in addressing a key unmet health equity need.^23^

Our finding of a wide variability in readmission rates amongst hospitals has been described before. There was a significant variability in patients’ outcomes across centers following HM3 LVAD implantation among individuals enrolled in the MOMENTUM 3 trial, with high-volume centers reporting better outcomes.^24^ This calls for models to define outcomes following LVAD and HT on a national scale incorporating center performance with inbuilt ways to provide feedback on areas for center-level improvement.^25^

As such, as discussed above, changes in the management of both LVAD and HT pertaining to anticoagulation, increasing GDMT use, and individualized immunosuppression have the potential to reduce readmission risks, cut healthcare costs and by doing so increase access to both definitive therapies in patients with advanced heart failure. In this light, there will be a need for continued assessment and comparative analysis of cost and resource utilization needed with HT and LVAD therapies on an ongoing basis in future.

### Limitations

Limitations inherent to observational, retrospective analyses, such as uncontrolled confounding, apply to this study. First, due to the administrative nature of the dataset used in this study, misclassification bias could be present because of improper selection of ICD codes. Second, the NRD does not contain data from outpatient appointments which could impact the readmission rates. Moreover, our data did not include visits to emergency departments or admissions to observation units, and therefore, we were not able to capture the true cost and burden on resource utilization. Additionally, excluding hospitalizations in October, November and December could have missed readmissions during these months. We were unable to examine the interaction between cost and resource utilization and race and ethnicity due to lack of information in NRD.

Further, on June 3, 2021, Medtronic announced the discontinuation of HVAD, meaning that HM3 is the sole remaining LVAD.^26,27^ Thus, our findings may partially reflect outcomes for a device no longer in production. While we have tried to capture all relevant outcomes, data extraction was based on billing codes, which may have limitations when applied to clinical analysis. Additionally, we were unable to comment on the costs of LVAD and donor organ acquisitions, as these were not recorded and could vary significantly between centers.

Despite these limitations, we do believe our study has several strengths. The national readmissions database captures information across large and diverse cross-sections of society across all socio-economic demographics, enhancing the external validity of our findings. Our study highlights the public health importance of these therapies for advanced heart failure and the potential ramifications of public health policy decisions on health equity. Finally, our study identifies specific areas where cost-effective improvements in treatment strategies could enhance patient care and outcomes.

## Conclusions

In this nationally representative dataset of the national readmission registry, heart transplants and implantation of durable left ventricular assist devices were both associated with significant short-term resource utilization and costs. Index hospitalization and re-admissions were higher for the LVAD group whereas HT was associated with higher re-admission-related costs. There remains a significant opportunity and need for modifying approach to post LVAD and HT care to reduce associated readmissions and costs of care.

## Data Availability

The data, analytic methods, and study materials are available through the HCUP website and may be used to reproduce the results. We also have added codes in supplementary table 1 for verification of analyses.

https://data.bls.gov/cgi-bin/cpicalc.pl

## Acknowledgements and source of funding

none

